# Multi-ancestry genome-wide analysis identifies effector genes and druggable pathways for coronary artery calcification

**DOI:** 10.1101/2022.05.02.22273844

**Authors:** Maryam Kavousi, Maxime M. Bos, Hanna J. Barnes, Christian L. Lino Cardenas, Doris Wong, the CHARGE Subclinical and Clinical Atherosclerosis Working Group, Christopher J. O’Donnell, Lawrence F. Bielak, Patricia A. Peyser, Rajeev Malhotra, Sander W. van der Laan, Clint L. Miller

**Affiliations:** Department of Epidemiology, Erasmus MC, University Medical Center Rotterdam, The Netherlands; Cardiovascular Research Center, Cardiology Division, Department of Medicine, Massachusetts General Hospital, Harvard Medical School, Boston, Massachusetts, USA; Department of Biochemistry and Molecular Genetics, University of Virginia, Charlottesville, Virginia, USA; Center for Public Health Genomics, University of Virginia, Charlottesville, Virginia, USA; Department of Medicine, Brigham and Women’s Hospital, Harvard Medical School, Boston, Massachusetts, USA; Cardiology Section, Department of Medicine, Veterans Affairs Boston Healthcare System, Boston, Massachusetts, USA; Department of Epidemiology, University of Michigan, Ann Arbor, Michigan, USA; Central Diagnostics Laboratory, Division Laboratories, Pharmacy, and Biomedical Genetics, University Medical Center Utrecht, Utrecht University, Utrecht, The Netherlands; Department of Public Health Sciences, University of Virginia, Charlottesville, Virginia, USA

## Abstract

Coronary artery calcification (CAC), a measure of subclinical atherosclerosis, predicts symptomatic coronary artery disease. Identifying genetic risk factors for CAC may point to new therapeutic avenues for preventing clinical disease. Here, we conducted a multi-ancestry genome-wide association study in 26,909 individuals of European ancestry and 8,867 individuals of African American ancestry. We identified 11 independent risk loci, of which 8 are novel for CAC. Some novel loci harbor candidate causal genes supported by multiple lines of functional evidence. Together, these findings help refine the genetic architecture of CAC, extend our understanding of the biological pathways underlying CAC formation, as well as identify druggable targets for CAC.

## Introduction

Coronary artery disease (CAD) is the leading cause of morbidity and mortality in developed countries^1,2^. Atherosclerosis is the primary etiology of CAD involving chronic lesion progression and luminal narrowing of arteries^3^. Subclinical atherosclerosis in the absence of clinical symptoms is associated with an increased risk of developing clinical CAD in both women and men from different ancestry groups independent of traditional risk factors^4–6^. Subclinical atherosclerosis can be detected noninvasively as coronary artery calcification (CAC) by cardiac computed tomography. Current clinical guidelines recommend assessment of CAC to clarify atherosclerotic cardiovascular disease (CVD) risk and to improve management decisions for those at borderline or intermediate atherosclerotic CVD risk^7^.

Based on family data, the estimated heritability for CAC is 30-40%^8,9^. Prior genome-wide association studies (GWAS) have identified non-coding single nucleotide polymorphisms (SNPs) at the 9p21 (*CDKN2B-AS1*) and 6p24 (*PHACTR1*) loci, as well as a protein-coding variant in *APOB* associated with a higher CAC quantity in individuals of European ancestry^10,11^. Another protein-coding variant in *APOE* was associated with CAC quantity in individuals of both European and African American ancestries^10^. These four identified genes for CAC are also associated with risk for CAD. Here, we carried out the largest CAC GWAS meta-analysis to date by analyzing 1000 Genomes imputed genotype data from 35,776 individuals of European and African ancestries through a collaboration between the Cohorts for Heart and Aging Research in Genomic Epidemiology (CHARGE) consortium^12^ and other participating cohorts. We then performed a series of downstream *in silico* functional genomic analyses to: 1) gain mechanistic and biological insights into how the identified genetic loci impact CAC quantity, 2) prioritize the most clinically relevant CAC loci, and 3) identify potential druggable targets for CAC. Notably, our *in vitro* and *ex vivo* experimental studies support our main genetic findings and provide motivation for future mechanistic studies.

## Results

### Multi-ancestry CAC genome-wide meta-analysis

We performed a meta-analysis by combining data from 35,776 individuals of European and African American ancestries. We identified 16 lead SNPs from 46 independent significant SNPs (r^2^ < 0.6; *P* < 5×10^−8^) in 11 independent genomic risk loci. Among these 11 loci, 8 are novel for CAC. The associations in *CDKN2B-AS1/CDKN2B* (9p21.3), *PHACTR1* (6p24.1), and *APOE* (19q13.32) replicated known findings^10,11^. We annotated the lead SNPs in the 11 loci and identified 2 missense lead SNPs in one novel gene and in *APOE*, and the remaining SNPs were annotated as non-coding. The specific *APOB* association reported earlier was not replicated here, most likely because the Old Order Amish were not included in this study and they have the highest frequency of this rare variant that is associated with CAC^13^.

### Refinement of identified CAC loci

We performed conditional analyses to identify secondary signals^14^, but did not identify new independent signals at these loci. We then performed credible set analyses to refine the signals at the 11 loci^15^. As expected, the 95% credible set reduced the number of candidate causal variants at the majority of loci, and even to a single candidate variant at loci *PHACTR1* and *APOE*. However, a considerable number of candidate SNPs remained at other loci.

We identified 38 candidate genes using the web-based platform FUMA^16^ through a combination of positional gene mapping, expression quantitative trait loci (eQTL) and chromatin interaction mapping. We identified another 2 candidate genes through a genome-wide gene association analysis (MAGMA)^17^. We also identified 3 candidate genes by annotating the nearest protein-coding genes that were not mapped through either of these methods. Taken together, we identified 43 candidate CAC genes.

Given the strong association between CAC and CAD, we further examined which of the CAC candidate genes are also associated with clinical CAD using public GWAS data^18,19^. Among our novel CAC loci, only one was nominally associated with CAD, while the others were not associated with clinical CAD. We observed similar results in our PheWAS analysis, where only one gene was significant after gene-based multiple testing correction, suggesting the other genes are likely CAC-specific.

**Table 1:** Significant associated loci and independent variants for CAC quantity in combined-ancestry meta-analysis.

| *rsID* | *Chr* | *Position (hg37)* | *Position (hg38)* | *Alleles* | *EAF* | *Effect* | *SE* | *P<sub>meta</sub>* | *I<sup>2</sup>* | *P<sub>I</sub><sup>2</sup>* | *Nearest gene* | *Annotation* |
| --- | --- | --- | --- | --- | --- | --- | --- | --- | --- | --- | --- | --- |
| **Known loci for coronary artery calcification quantity** |  |  |  |  |  |  |  |  |  |  |  |  |
| *rs10456561* | 6 | 12,887,465 | 12,887,233 | A/G | 0.036 | 0.375 | 0.069 | 4.76x10<sup>-8</sup> | 14.7 | 0.292 | *PHACTR1* | intronic |
| *rs35355695* | 6 | 12,891,103 | 12,890,871 | G/T | 0.256 | -0.115 | 0.020 | 4.87x10<sup>-9</sup> | 0 | 0.683 | *PHACTR1* | intronic |
| *rs9349379* | 6 | 12,903,957 | 12,903,725 | A/G | 0.623 | -0.218 | 0.020 | 6.11x10<sup>-29</sup> | 0.5 | 0.453 | *PHACTR1* | intronic |
| *rs10811650* | 9 | 22,067,593 | 22,067,594 | A/G | 0.619 | -0.195 | 0.018 | 2.15x10<sup>-27</sup> | 0 | 0.744 | *CDKN2B-AS1* | ncRNA intronic |
| *rs72652478* | 9 | 22,102,043 | 22,102,044 | C/G | 0.957 | -0.479 | 0.081 | 3.82x10<sup>-9</sup> | 0 | 3.491 | *CDKN2B-AS1* | ncRNA intronic |
| *rs62555371* | 9 | 22,107,238 | 22,107,239 | A/T | 0.866 | 0.270 | 0.032 | 5.49x10<sup>-17</sup> | 4 | 0.407 | *CDKN2B-AS1* | ncRNA intronic |
| *rs4977575* | 9 | 22,124,744 | 22,124,745 | C/G | 0.455 | -0.264 | 0.018 | 7.49x10<sup>-47</sup> | 5.2 | 0.390 | *CDKN2B-AS1 (dist.: -3,651 bp),*<br>*CDKN2B (dist: 115,440 bp)* | intergenic |
| *rs7412* | 19 | 45,412,079 | 44,908,822 | C/T | 0.090 | -0.313 | 0.039 | 4.42x10<sup>-16</sup> | 0 | 0.940 | *APOE* | missense |

Top lead SNP in genomic risk loci associated with coronary artery calcification quantity at a significance level of $P < 5 \times 10^{-8}$ , for the combined-ancestry meta-analysis (up to 35,776 individuals from 22 studies). SNP effect sizes (Effect) and two-sided P-values ( $P_{\text{META}}$ ) were derived from a meta-analysis using a fixed-effects model with an inverse-variance weighted approach. rsID: the dbSNP number of the lead SNP. The chromosomal (*Chr*) position is given in hg37 and hg38. Alleles: effect and other allele. EAF: effect allele frequency. SE: standard error of the effect. $I^2$ : heterogeneity statistic. $P_I^2$ : the p-value of the heterogeneity statistic.

### Prioritization of candidate causal genes and variants

To prioritize candidate causal genes and variants we performed Summary-based Mendelian Randomization (SMR)^20^ and colocalization^21^. By integrating the European ancestry CAC meta-analysis summary statistics and cardiometabolic tissue *cis*-eQTLs from STARNET^22,23^, we identified 11 and 18 gene-trait associations using mapped eQTLs in atherosclerotic aortic root (AOR) and internal mammary artery (MAM) tissues, respectively. To provide additional functional fine-mapping evidence at the variant-level, we performed colocalization using coloc^21^, which revealed colocalization of CAC variants with cis-eQTLs in 22, 25, and 7 genes, in AOR, MAM, and liver (LIV), respectively. We observed the strongest evidence of colocalization at known CAD loci, *PHACTR1* in AOR, consistent with recent fine-mapping studies^24^. As expected we observed substantial overlap between prioritized genes associated with CAC and CAD traits (PP4 >0.80 for both traits). However, we also identified a subset of genes with strong evidence of colocalization with CAC (PP4 >0.80) but not CAD (PP4 <0.50). Interestingly, one novel locus was identified as the target gene using SMR in both AOR and MAM, which may suggest distinct mechanisms influencing CAC risk that are independent of CAD.

To further resolve the regulatory mechanisms of GWAS variants^25,26^, we performed epigenomic based fine-mapping analyses using the activity-by-contact (ABC) ^27^ and enhancer-gene linking^28^ methods. Using a suggestive threshold for CAC associated variants (*P* < 1 × 10^−5^), we identified 42 and 54 variants overlapping enhancer-promoter contacts for predicted target genes in human coronary artery smooth muscle cells (HCASMC) and coronary artery, respectively). Notably, this provided additional support for novel CAC variants regulating several novel loci, and confirmed a regulatory variant in the 9p21 locus (rs1537373) affecting *CDKN2B*. We confirmed these findings using our recent single-nucleus ATAC-seq dataset in healthy and diseased coronary arteries^29^, which identified credible CAC variants at several novel loci overlapping cell type-specific peak-to-gene links. These results demonstrate that several identified CAC GWAS signals map to relevant genes in the vascular wall providing candidate regulatory mechanisms for the CAC associations.

### Identification of target pathways, cell types, and plaque phenotypes

Using gene-set pathway enrichment analysis for the candidate CAC genes, we identified significant evidence for several relevant pathways.

We investigated the overall expression profiles for the candidate CAC genes in bulk GTEx tissues. Many of the candidate genes demonstrated expression in artery tissues and clustered together, suggesting similar cellular profiles. We also explored the cellular distribution in single-cell gene expression data from atherosclerotic coronary artery^30–32^ and carotid plaques^33^.

To gain insight in the pathobiology of the 16 lead CAC SNPs, we assessed their associations with plaque morphology. We examined 7 plaque morphological characteristics measured in carotid advanced plaques^34^. *APOE* was associated with increased intraplaque fat content and vessel density, and decreased collagen content which are known features of increased plaque vulnerability. Individual variant analyses at the remaining loci were most significant for plaque calcification (*PHACTR1*), smooth muscle cell (*CDKN2B-AS1*) and intraplaque neovessel density (*PHACTR1*).

### Heritability, genetic correlations, and Mendelian randomization

We applied linkage disequilibrium score regression (LDSC)^35^ to estimate the heritability of CAC in those of European ancestry and observed that the genome-wide set of variants account for 16% (s.e.= 2.5%) of the variance in CAC. This estimate represents almost half of the percent heritability estimated from phenotypic correlations among relatives.

We estimated the genetic correlation between CAC and other measures of subclinical atherosclerosis, clinical CVD, family history of CVD, and CVD risk factors in those of European ancestry. There were significant genetic correlations between CAC and carotid plaque and abdominal aortic calcification as well as several clinical outcomes including CAD and myocardial infarction. CAC had significant genetic correlations with high cholesterol, using cholesterol lowering medication, hypertension, body mass index (BMI), waist circumference, and whole-body fat mass.

**Figure 1:**
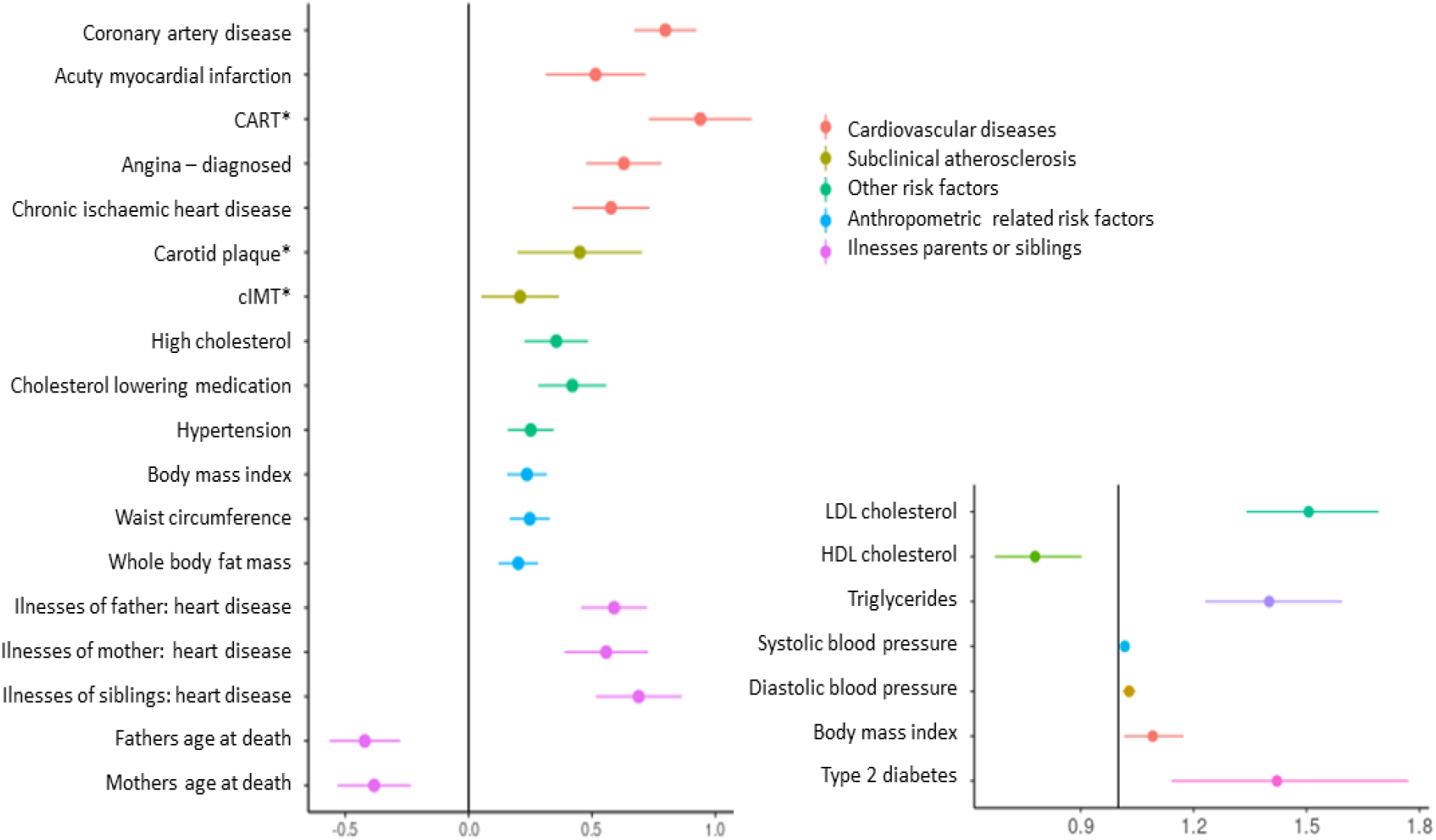
Genetic correlations for CAC and Mendelian randomization for cardiovascular disease risk factors. *Left:* Genetic correlation of CAC quantity with cardiovascular disease risk factors, subclinical and clinical cardiovascular disease. *Right:* Mendelian randomization results for CAC quantity and cardiovascular disease risk factors.

We then performed Mendelian Randomization (MR) analyses to assess potential causality of CVD risk factors with CAC. Low-density lipoprotein (LDL) cholesterol, triglycerides, systolic and diastolic blood pressure, BMI, and risk for type 2 diabetes were causally associated with an increase in CAC quantity, while an increase in high-density lipoprotein (HDL) cholesterol was causally associated with a decrease in CAC quantity. Moreover, CAC quantity was associated with clinical CAD. In the sensitivity analyses, all associations showed effect estimates in similar directions thereby providing evidence for a robust association. Further, MR Egger regression analyses revealed no evidence for horizontal pleiotropy.

### Druggability analysis

To investigate the potential clinical utility of CAC candidate genes at the 11 independent risk loci, we performed an integrative druggability analysis. Three loci were identified as targets of clinically actionable compounds, which have potential for repurposing approved drugs and informing clinical trials for CAC. Notably, several of the CAC-specific genes represent targets of the druggable genome, which should be considered for preclinical studies of CAC. By querying the drug-gene interaction database (DGIdb) and DrugBank database, we revealed approved compounds under investigation targeting other novel loci. These findings offer preclinical, repurposing and lifestyle modification opportunities via these targets for both subclinical atherosclerosis and clinical CAD involving vascular calcification^36^. Notably these target genes were also supported by multiple lines of statistical and functional fine-mapping evidence and were validated by our *in vitro* and *ex vivo* validation studies.

## Discussion

We identified 16 independent genome-wide significant variant associations for CAC quantity at 11 distinct genomic loci by leveraging genome-wide data on 35,776 participants, including 8 loci not previously reported to be associated with CAC. Through integrating functional data with GWAS results as well as gene-based analysis, we identified 43 candidate genes for CAC quantity. Some of the identified genes are implicated in relevant pathways. Notably, several of our CAC genes represent identified druggable targets revealing opportunities for either drug repurposing or development strategies.

As the hallmark of atherosclerosis, coronary artery calcification is strongly associated with future coronary events. Consistent with the shared etiology between CAC and CAD, several of our novel CAC genes were previously reported in association with CAD. Our colocalization analyses also indicated substantial overlap between prioritized genes in CAC and CAD. There is debate as to whether coronary calcification is a consequence (‘scar tissue’) of the atherosclerotic process or is causally related to CAD^37,38^. Our MR analyses provided evidence for a causal association between CAC quantity with CAD events. However, several of our CAC genes have so far not been reported to be associated with CAD. In line, these CAC-specific genes showed strong evidence for colocalization with CAC but not with CAD. The relationship of CAC to plaque instability is complex and incompletely understood^39^. While higher CAC quantity correlates with overall atherosclerotic plaque burden and luminal stenosis, CAC is not a marker of plaque vulnerability. It is suggested that stable, slowly progressive large plaques, leading to a positive remodeling of the vessel, do not readily correlate with onset of symptoms and clinical CAD events. In contrast, unstable plaques, at high risk of producing symptomatic rupture, carry highly inflamed fibrous caps and are not necessarily more stenotic^40^.

CAC reflects the vessel’s memory of lifetime exposure to risk factors. While observational studies suggest a role for hypertension, higher BMI, and type 2 diabetes mellitus in development and progression of CAC, evidence regarding the association of unfavorable lipid profile with coronary calcification, in particular progression of CAC remains unclear^41,42^. Moreover, important questions remain about causality of these risk factors. We found evidence of significant genetic correlations between high cholesterol, using cholesterol lowering medication, hypertension, BMI, waist circumference, and whole body fat mass with CAC quantity. Our MR analyses further provided evidence for causal association between modifiable cardiovascular risk factors, including LDL and HDL cholesterol, triglycerides, type 2 diabetes mellitus, and BMI with CAC quantity. These findings emphasize the value of optimal risk factor control for reduction of atherosclerosis burden and further extend our knowledge of the pathways underlying coronary calcification and may inform new therapeutic strategies.

### Limitations and strengths

We included only individuals of European and African-American ancestry in our meta-analyses. Discovery of additional novel CAC loci will require participants from other ancestral populations. With the ever-increasing size of CAD GWASs, it is also anticipated that some of our CAC-specific loci will be associated with CAD over time. Also, we used the 1000 Genomes reference imputation panel which might limit the ability to uncover less common variants. Future multi-ancestry meta-analyses will benefit from using larger, more diverse reference imputation panels such as TOPMed^43^. Our eQTL-based fine-mapping methods assume that non-coding variants primarily act through the regulation of gene expression, however it is known that many variants also function through splicing, translation, and other modes of molecular regulation^44^. Future integration with multi-omics QTLs in healthy and diseased tissues and stimulated cell types (e.g. osteogenic conditions) may resolve additional mechanisms for these associations^45,46^.

There are many strengths to our study which employed a series of complementary statistical, functional fine-mapping, and *in vitro* and *ex vivo* experimental studies. First we employed an array of post-GWAS fine-mapping methods using both publicly available datasets and those from disease-specific cohorts. While many earlier studies have relied on transcriptome datasets from deceased donors to perform fine-mapping (e.g. those in GTEx), these may not reflect the genes altered in specific cell types in subclinical atherosclerosis. To mitigate this, we utilized the STARNET cohort of eQTLs, which were derived from living subjects during coronary artery bypass graft surgical procedures. We also annotated genes by leveraging unique coronary and carotid artery atherosclerosis tissue biobanks, which provided more relevant context to the affected plaque phenotypes and cell types. Finally, we employed druggability analyses and *in vitro* and *ex vivo* functional validation assays to help inform translational strategies and identify cell-specific mechanisms for these candidate targets.

In conclusion, we discovered 8 novel loci associated with CAC. Extensive post-GWAS fine-mapping and annotation provided evidence for cell- and disease-specific expression of the CAC-specific genes in coronary and carotid arterial plaque tissue. Importantly, many of these genes encode for proteins that were identified as predicted targets of approved drugs or investigational compounds. Future studies should elucidate the molecular mechanisms of these genes in the cells of the arterial wall, preclinical testing of candidate genes, and focus on creating a saturated map of common and rare variants for CAC through the inclusion of diverse ancestries.

## Methods

### Ethics statement

All human research was approved by the relevant institutional review boards for each study and conducted according to the Declaration of Helsinki. All participants provided written informed consent. The details per cohort are given in **Supplemental Material Table 1**.

### Study populations and CAC assessment

The GWAS for CAC quantity included 16 different cohorts. These cohorts contributed 26,909 participants of European ancestry and 8,867 participants of African American ancestry. All cohorts followed standardized protocols for the ascertainment of CAC quantity and statistical analyses. CAC was evaluated using computed tomography. We used data from the baseline examination or the first examination in which CAC was assessed.

### Genotyping, imputation, and study-level quality control

Association analyses were performed using standardized protocols. Within each study, linear regression was used to model CAC quantity with an additive genetic model (SNP dosage) adjusted for age, sex, and up to 10 principal components. Extensive quality control (QC) was applied to the data. Each study conducted genome-wide imputation using a Phase 1 integrated (March 2012 release) reference panel from the 1000G Consortium using IMPUTE or MaCH/minimac and used Human Reference Genome Build 37. There was little evidence for population stratification in any studies. Sample QC was performed with exclusions based on call rates, extreme heterozygosity, sex discordance, cryptic relatedness, and outlying ancestry. SNP QC excluded variants based on call rates across samples and extreme deviation from Hardy–Weinberg equilibrium. Non-autosomal SNPs were excluded from imputation and association analysis. We used the EasyQC R package (v23.8) to perform QC for each study before the meta-analysis and excluded markers absent in the 1000G reference panel: no-A/C/G/T/D/I markers; duplicate markers with low call rate; monomorphic SNPs and those with missing values in alleles, allele frequency, and/or beta estimates; SNPs with large effect estimates or standard error (SE) ≥10; and SNPs with allele frequency difference >0.3 compared to 1000G reference panel.

### Meta-analysis

Since the joint analysis is more efficient than a two-stage discovery-replication analysis we chose to perform a meta-analysis on all available samples rather than use a two-stage discovery-replication strategy^47^. A meta-analysis of GWAS was performed using fixed-effects meta-analysis METAL, using SNP P-values weighted by sample size. Summary statistics from each study were combined using an inverse variance weighted meta-analysis. Additional filters were applied during meta-analyses including imputation quality (MACH r^2^ < 0.3 and IMPUTE info <0.4), a minor allele frequency (MAF) <0.01, and SNPs that were not present in at least four studies or in both ancestry groups. Moreover, the variants with heterogeneity I^2^ ≥70% in the meta-analysis were excluded, leaving 8,586,047 variants. The genome-wide significance threshold was considered at *P* < 5.0 × 10^−8^.

### Genomic risk loci definition

We used FUMA version v1.3.6a (see URLs) to obtain the genomic risk loci and functional information for the relevant SNPs in these loci. FUMA combines several external data sources to provide comprehensive annotation information. First, independent significant SNPs, at *P* < 5 × 10^−8^ and at r^2^ < 0.6 were identified. We further defined the lead SNPs as a subset of the independent significant SNPs that were in approximate LD with each other at r^2^ < 0.1. Physical regions in LD with these lead SNPs that were > 250 kilobases (kb) apart from each other were identified as independent genomic risk loci. We defined the genomic risk loci by identifying all SNPs in LD (r^2^ ≥ 0.6) with one of the independent significant SNPs; the region containing all these candidate SNPs was considered to be a single independent genomic risk locus. In naming the nearest genes for the independent loci, we mapped the nearest gene (protein coding or non-coding) to the lead SNPs based on physical location to the transcription start sites (TSS). We also assigned the nearest protein-coding genes at these loci using canonical TSS from GENCODE (v30) in ANNOVAR functional annotator version 2019-10-24^48^.

### Functional annotation and gene mapping

We first annotated the function of variants using the Ensembl Variant Effect Predictor (v92)^49^. We used FUMA (v1.3.6a, http://fuma.ctglab.nl/)^16^ for functional annotation of the GWAS results based on 1000G phase 3 (version 5 based on the EUR populations) and Ensembl 92 (only protein coding genes were considered, n = 19,177). Variants were filtered while performing annotation based on positional gene mapping (maximum distance 10kb), eQTL-based gene mapping, and 3D chromatin interaction mapping using a minimum CADD score ≥ 12.37 (considered to be suggestively deleterious), a maximum RegulomeDB score of 7, a maximum 15-core chromatin state of 7 using all available tissues. For eQTL mapping aorta, coronary and tibial artery derived data (GTEx v8) were used and a FDR < 0.05 was applied. For the 3D chromatin interaction analysis a FDR < 10^−6^ threshold was applied using all the available data from Roadmap. We defined the promoter region in a window of 250 Kb upstream and 500 Kb downstream of the transcription start site (TSS) to overlap genes with significantly interacting regions, and only variants overlapping enhancers and genes whose promoters overlap epigenomic regions were used. Variants were mapped against the GWAS Catalog version e104_r2021-09-15^50^ and ANNOVAR version 2017-07-1^48^ as implemented in FUMA.

### Gene-based test/gene-set analysis

Gene-based test and gene-set analysis are methods that enable us to summarize SNP associations at the gene level and associate the set of genes to biological pathways. We used MAGMA v1.08^17^ through FUMA to perform gene-based analyses using the summary statistics of the combined meta-analysis in a window of 50kb around 19,177 protein coding genes (mapped to GRCh37/hg19 based on Ensembl 92). We used default settings to calculate empirical p-values derived from 1,000 permutations and set a nominal p-value conservatively at 0.05/19,177 = 2.61×10^−6^. MAGMA leverages the per-variant test statistics by applying a multiple regression model to derive an empirical p-value for association of individual genes considering the linkage disequilibrium structure that exists among variants and potential multi-marker effects^17^.

### Gene and variant level fine-mapping

In order to further prioritize genes at CAC loci, we used two colocalization methods, summary level mendelian randomization (SMR)^20^ and coloc^21^. We integrated the CAC GWAS data with gene expression data from the STARNET cohort of 7 cardiometabolic tissues: atherosclerotic aortic root (AOR), whole blood (Blood), liver (LIV), mammary artery (MAM), subcutaneous fat (SF), visceral fat (VF), and skeletal muscle (SKLM), derived from ∼600 individuals of European ancestry^22^, with a focus on AOR, MAM and LIV tissues. We first performed an SMR analysis to test whether top CAC GWAS variants influence the phenotype through perturbation of gene expression in these atherosclerosis relevant tissues. We considered only genes with at least 1 cis-eQTL *P* < 5×10^−5^ for colocalization. To account for a model of linkage, where two distinct signals drive the association with gene expression and CAC, we used the 1000G European LD reference panel and the post-hoc post-hoc heterogeneity in dependent instruments (HEIDI) test 1 to exclude loci with evidence of linkage or heterogeneity in the genetic instruments. We performed the SMR/HEIDI test on 5233 and 5293 eGenes (peQTL < 5E^-5^) in AOR and MAM tissues, respectively, to identify genes that passed a q-value^51^ threshold <0.10 (AOR: PSMR <3E^-4^, MAM: *P*_SMR_ <4E^-4^) and HEIDI test (*P*_HEIDI_ >0.01).

We also performed a colocalization analysis using the R based package, coloc^21^. This Bayesian statistical approach calculates the posterior probability that the CAC GWAS and eQTL data from different STARNET tissues share a common signal under the one causal variant assumption. Following a filtering step to include only eQTLs with p-value <0.05, we tested for colocalization between overlapping variants in the CAC GWAS and STARNET expression data. We considered PP4 >0.80 as strong evidence of colocalization. We considered strongly colocalized loci for CAC as those having PP4 >0.8 for CAC but PP4 <0.5 for CAD, and strongly shared colocalized loci as those having PP4 >0.8 for both traits.

### Pathway enrichment analysis

Gene-based association analyses were performed using MAGMA^17^ using both EA and AA CAC summary statistics (both unfiltered and subset to P<1E^-05^). Gene-based p-values were computed based on the total sample size, and a gene annotation window of 2 or 5kb upstream and 1kb downstream of candidate genes. GTEx v8 blood vessel tissues (coronary, aorta, and tibial artery) were used for the MAGMA gene expression analysis. We also imported CAC strongly colocalized and CAC/CAD shared colocalized gene sets into enrichR (2020 update) to determine enriched pathways using a combination of databases including BioPlanet, BioCarta, KEGG, WikiPathways, Reactome, and PANTHER^52^.

### Activity-by contact and enhancer linking

We used the combined EA and AA CAC variants filtered to suggestive associations (P<1E^-05^) to intersect with genome-wide enhancer-gene predictions calculated in the activity-by-contact models^27^. We used both the ENCODE coronary artery tissue and human coronary artery smooth muscle cell based annotations from H3K27ac histone modification ChIP-seq and chromatin accessibility datasets, and we considered significant enhancer-gene predictions with ABC scores >0.02, as previously described^53^. For enhancer-gene linking annotations we again used the above CAC summary associations at P<1.0 × 10^−5^ and intersected these variants with human aortic enhancer states (6,7, and 12) calculated from the Epigenomics Roadmap consortium^28^.

### snATAC-seq analysis

Atherosclerotic coronary artery segments were obtained from explanted hearts from 41 patients undergoing heart transplantation or donor hearts procured for research purposes at Stanford University. All samples were collected under Institutional Review Board (IRB) approval and written informed consent. Frozen tissues were transferred to the University of Virginia through a material transfer agreement and IRB approved protocols and stored at -80 C until day-of-processing. For snATACseq analysis of human coronary artery samples, nuclei were isolated from frozen tissues, purified over an Opti-Prep sucrose gradient as described^29^ and subjected to 10X Genomics based library preparation and sequencing on a NovaSeq 6000 (paired end, 2 × 50 bp) to achieve ∼50,000 unique fragments per cell. Initial pre-processing was performed using the 10X Genomics pipeline (Cell Ranger ATAC v1.2.0). All ATAC-seq reads were mapped to the human genome reference hg38 build using the default parameters. Approximately 28,000 cells were included in the clustering analysis after filtering for high-quality cells with transcription start site (TSS) enrichment >7.0 and >10,000 fragments using the ArchR package (v.1.0.1) package^54^. ArchR was also used for downstream analyses including dimensionality reduction, clustering, calculation of imputed gene scores, and Peak2gene links as described^29^. snATAC tracks were visualized on the UCSC browser and compared with existing bulk coronary artery ATAC-seq tracks.

### GTEx analysis

Candidate CAC genes were queried in the Genotype Tissue Expression (GTEx) database (version 8) via the online portal (https://gtexportal.org/home/). This release includes 54 unique human tissues from 948 donors. Bulk RNA-seq processing and analysis details are available on the portal website. Queries of normalized mean gene expression levels per tissue were limited to the most relevant tissues and clustered using the Multi-Gene Query Tool.

### Carotid plaque analyses

Atherosclerotic plaques were obtained from patients undergoing a carotid endarterectomy (CEA) procedure and included in the Athero-Express Biobank Study (AE, www.atheroexpress.nl, approved and registered under number TME/C-01.18), an ongoing biobank study in Utrecht, The Netherlands^55^. The study design of the AE was described before^55^, but in brief: during surgery blood and plaques are obtained, stored at -80°C and plaque material is routinely used for standardized (immuno)histochemical analysis^55,56^.

We quantitatively scored the number of macrophages (CD68) and smooth muscle cells (SMCs, α-actin), as percentage of the microscope field area by computerized analysis. Intraplaque vessel density (CD34) was assessed as the average number per 3 hotspots. Intraplaque hemorrhage (IPH) was scored as no/yes using a hematoxylin and eosin staining (HE). Intraplaque fat was defined as less or more than 40% fat per total plaque area using HE. The amount of calcification (using HE) and collagen (picrosirius red) were binary scored as no/minor vs. moderate/heavy staining. Assessment of overall plaque vulnerability was performed as previously described by Verhoeven et al.^57^. All histological observations were performed by the same dedicated technician and interobserver analyses have been reported previously^58^.

For the genetic analyses of these histological plaque morphological characteristics, we isolated DNA following standardized protocols as described before^34^. In short, the AE study was genotyped in three separate, but consecutive experiments and carried out according to OECD standards: Athero-Express Genomics Study 1 (AEGS1) including 891 patients, the Athero-Express Genomics Study 2 (AEGS2) including 954 patients, and the Athero-Express Genomics Study 3 (AEGS3) including 658 patients. We adhered to community standard quality control and assurance (QCA) procedures of the genotype data as described before^34,59^. After QCA these comprise 890 samples and 407,712 SNPs in AEGS1, 869 samples and 534,508 SNPs in AEGS2, and 649,954 samples and 534,508 SNPs in AEGS3 remained. Missing genotypes were imputed with 1000G phase 3, version 5 and HRC release 1.1 as a reference using the Michigan Imputation Server^60^. These results were further integrated using QCTOOL v2, where HRC imputed variants are given precedence over 1000G phase 3 imputed variants.

For the association of the 11 independent loci with the plaque vulnerability and plaque morphological characteristics we queried 1000G phase 3 using FUMA and included 853 variants ±250kb with LD r^2^ surrounding the top loci. Next we performed regression analyses adjusted for age, sex and principal components, thus: phenotype ∼ age + sex + chip-used + PC1 + PC2 + year-of-surgery. Continuous variables were inverse-rank normal transformed.

More details on the single-cell RNA sequencing methods of the Athero-Express Biobank Study are given in the Supplementary Material, in short we collected carotid plaques from 35 individuals, isolated and processed RNA as described before^61^, while employed the CEL-seq2 method^62^.

### Heritability estimation of CAC

We used LD-score regression^35^ to estimate the proportion of variance in CAC that could be explained by the aggregated effect of the SNPs in those of European ancestry. The method assumes that an estimated SNP effect includes effects of all SNPs in LD with that SNP. On average, a SNP that tags many other SNPs will have a higher probability of tagging a causal variant than a SNP that tags few other SNPs.

Thus, SNPs with a higher LD-score have, on average, stronger effect sizes than SNPs with lower LD-scores. By regressing the effect size obtained from the GWAS against the LD-score for each SNP, the slope of the regression line will provide an estimate of heritability based on the analyzed SNPs. We included 1,167,424 SNPs with available betas. After merging with the European reference panel, 1,164,129 SNPs remained. SNP heritability was estimated using European LD scores from the 1000 Genomes Project Phase 3 data for the HapMap3 SNPs, downloaded from https://data.broadinstitute.org/alkesgroup/.

### Genetic correlations

We used cross-trait LD-score regression to estimate the genetic covariation between traits using GWAS summary statistics^63^. The genetic covariance is estimated using the slope from the regression of the product of *z*-scores from two GWAS studies on the LD-score. The estimate obtained from this method represents the genetic correlation between the two traits based on all polygenic effects captured by SNPs. Standard LD-scores were used as provided by Bulik-Sullivan et al. based on the 1000 Genomes reference set^64^, restricted to European ancestry populations.

### Mendelian randomization analyses

Two-sample Mendelian randomization (MR) using summary-level data was applied to investigate potential causality between CVD risk factors and a higher CAC quantity^65^, and a higher CAC quantity with CAD. We selected SNPs associated with each CVD risk factor at the genome-wide level of significance (*P* < 5×10^−8^) as our exposure. SNPs were identified from publically available genome-wide association studies and only studies with European ancestry were considered. We excluded SNPs that had LD with other variants, were absent from the LD reference panel or were palindromic with intermediate allele frequencies. A total of 75 independent genetic instruments for LDL cholesterol, 86 for HDL cholesterol^66^, 54 for triglycerides^66^, 428 for low systolic blood pressure^67^, 428 for diastolic blood pressure^67^, 490 for body mass index^68^ and 118 for type II diabetes were included. As an outcome, the summary statistics of our CAC GWAS were used. For the association of CAC with CAD, we selected the lead 16 independent SNPs associated with CAC quantity in the current GWAS as the exposure and summary statistics of CAD as our outcome^19^. Inverse variance-weighted (IVW) analyses were used in which combined effects of the individual genetic instruments on the outcome, here being CAC quantity, result in a weighted mean estimate of a genetically determined increase in exposure on the outcome^69^. Moreover, we performed weighted median estimator (WME) and MR Egger regression analyses as sensitivity analyses to rule out potential bias caused by directional pleiotropy^70^. The analyses and data visualizations were performed using MRCIEU/TwoSampleMR^71^ and ggplot2.

### PheWas

For our PheWAS analysis we queried selected CAC genes based on their multiple lines of functional evidence and specific association with CAC. The selected genes were queried in the GWAS ATLAS resource (https://atlas.ctglab.nl)^72^ and the TOPMed PheWeb (https://pheweb.org/UKB-TOPMed/pheno/411.4).

### Druggability analysis

We used the DGIDB database to import CAC strongly colocalized or CAC/CAD shared colocalized gene sets to determine the potentially druggability of the candidate gene targets. We imported these gene sets into the Protein Data Bank (PDB) to query the pocket druggability scores using the PockDrug prediction models. We queried protein targets for available active ligands in ChEMBL. We queried gene targets in the druggable genome using the most recent druggable genome list established from the NIH Illuminating the Druggable Genome Project (https://github.com/druggablegenome/IDGTargets) also available through the Pharos web platform. We also queried FDA-approved drugs, late-stage clinical trials and disease indications in the DrugBank and ClinicalTrials.gov databases and provided results for CVD-relevant indications.

## Data Availability

All data produced in the present study are available upon request to the authors.

