## Supplemental Material for "Multi-ancestry genome-wide analysis identifies effector genes and druggable pathways for coronary artery calcification"

**Table of Contents**

**Supplemental Tables**	**3**

### Supplemental Tables

**Supplemental Table 1: List of included cohorts and their respective Ethical Review Boards’ (ERB) or Institutional Review Boards' (IRB) decisions.**

| **Study (acronym)** | **Study (full name)** | **Ancestry** | **ERB/IRB description** | **Decision** | **Reference #1** | **Reference #2** | **Reference #3** | **Reference #4** |
| --- | --- | --- | --- | --- | --- | --- | --- | --- |
| ***AGES*** | Age, Gene, Environment, Susceptibility Study | European Ancestry | National Bioethics Committee in Iceland that acts as the institutional review board for the Icelandic Heart Association and by the National Institute on Aging Intramural Institutional Review Board. Iceland. | Approved | <https://www.ncbi.nlm.nih.gov/pmc/articles/PMC7665790/> |  |  |  |
| ***ASPS*** | Austrian Stroke Prevention Study | European Ancestry | Medical University of Graz, Austria. | Approved | <https://dx.doi.org/10.1016%2Fj.clineuro.2012.12.016> |  |  |  |
| ***CARDIA*** | Coronary Artery Risk Development in Young Adults Study | European Ancestry | Four Field Centers Institutions were part of CARDIA: 1 University of Alabama at Birmingham, Ala, 2 Northwestern University, Chicago, Ill., 3 University of Minnesota, Minneapolis, Minn., and 4 Kaiser Permanente Medical Care Program, Oakland, Calif. Institutional Review Board IRB approvals were obtained from each field center institution. Each subject signed an informed consent document. USA. | Approved | <https://clinicaltrials.gov/ct2/show/NCT00005130> | <https://doi.org/10.1161/JAHA.118.010586> | <https://www.cardia.dopm.uab.edu/contact-cardia> | <https://doi.org/10.1016/0895-4356(88)90080-7> |
| ***CARDIA*** | Coronary Artery Risk Development in Young Adults Study | African American | Four Field Centers Institutions were part of CARDIA: 1 University of Alabama at Birmingham, Ala, 2 Northwestern University, Chicago, Ill., 3 University of Minnesota, Minneapolis, Minn., and 4 Kaiser Permanente Medical Care Program, Oakland, Calif. Institutional Review Board IRB approvals were obtained from each field center institution. Each subject signed an informed consent document. USA. | Approved | <https://clinicaltrials.gov/ct2/show/NCT00005130> | <https://doi.org/10.1161/JAHA.118.010586> | <https://www.cardia.dopm.uab.edu/contact-cardia> | <https://doi.org/10.1016/0895-4356(88)90080-7> |
| ***COPDGene*** | Examining the Genetic Factors That May Cause Chronic Obstructive Pulmonary Disease Study | European Ancestry | Twenty one clinical study centers throughout the United States enrolled participants. Each study site obtained local Institutional Review Boards IRB approval to enroll participants and all subjects provide informed consent to participate in the study. A Certificate of Confidentiality from the US Department of Health and Human Services was obtained at the onset of the study to provide additional protection for the research participants and their subsequently generated data on genetic markers. The participating centers were: Ann Arbor VA, Ann Arbor, MI, Baylor College of Medicine, Houston, TX, Brigham and Women's Hospital, Boston, MA, Columbia University, New York, NY, Duke University Medical Center, Durham, NC, Fallon Clinic, Worcester, MA, Health Partners Research Foundation, Minneapolis, MN, Johns Hopkins University, Baltimore, MD, Los Angeles Biomedical Research Institute at Harbor UCLA Medical Center, Los Angeles, CA, Michael E. DeBakey VAMC, Houston, TX, Minneapolis VA, Minneapolis, MN, Morehouse School of Medicine, Atlanta, GA, National Jewish Health, Denver, CO, Temple University, Philadelphia, PA, University of Alabama, Birmingham, AL, University of California, San Diego, CA, University of Iowa, Iowa City, IA, University of Michigan, Ann Arbor, MI, University of Minnesota, Minneapolis, MN, University of Pittsburgh, Pittsburgh, PA, University of Texas Health Science Center at San Antonio, San Antonio, TX. USA. | Approved | <https://dx.doi.org/10.1186%2Fs12931-019-1237-1> | <https://clinicaltrials.gov/ct2/show/NCT00608764> |  |  |
| ***COPDGene*** | Examining the Genetic Factors That May Cause Chronic Obstructive Pulmonary Disease Study | African American | Twenty one clinical study centers throughout the United States enrolled participants. Each study site obtained local Institutional Review Boards IRB approval to enroll participants and all subjects provide informed consent to participate in the study. A Certificate of Confidentiality from the US Department of Health and Human Services was obtained at the onset of the study to provide additional protection for the research participants and their subsequently generated data on genetic markers. The participating centers were: Ann Arbor VA, Ann Arbor, MI, Baylor College of Medicine, Houston, TX, Brigham and Women's Hospital, Boston, MA, Columbia University, New York, NY, Duke University Medical Center, Durham, NC, Fallon Clinic, Worcester, MA, Health Partners Research Foundation, Minneapolis, MN, Johns Hopkins University, Baltimore, MD, Los Angeles Biomedical Research Institute at Harbor UCLA Medical Center, Los Angeles, CA, Michael E. DeBakey VAMC, Houston, TX, Minneapolis VA, Minneapolis, MN, Morehouse School of Medicine, Atlanta, GA, National Jewish Health, Denver, CO, Temple University, Philadelphia, PA, University of Alabama, Birmingham, AL, University of California, San Diego, CA, University of Iowa, Iowa City, IA, University of Michigan, Ann Arbor, MI, University of Minnesota, Minneapolis, MN, University of Pittsburgh, Pittsburgh, PA, University of Texas Health Science Center at San Antonio, San Antonio, TX. USA. | Approved | <https://dx.doi.org/10.1186%2Fs12931-019-1237-1> | <https://clinicaltrials.gov/ct2/show/NCT00608764> |  |  |
| ***DHS*** | Diabetes Heart Study | European Ancestry | Institutional Review Board at the Wake Forest University School of Medicine (WFUSM) and all participants gave written informed consent. Winston-Salem, NC, USA. | Approved | <https://pubmed.ncbi.nlm.nih.gov/21409311/> | <https://link.springer.com/article/10.1007/s00125-005-0017-2> |  |  |
| ***FamHS*** | Family Heart Study | European Ancestry | Washington University School of Medicine IRB. St. Louis, MO, USA. | Approved | <https://www.ncbi.nlm.nih.gov/pmc/articles/PMC7665790/> |  |  |  |
| ***FamHS*** | Family Heart Study | African American | Washington University School of Medicine IRB. St. Louis, MO, USA. | Approved | <https://www.ncbi.nlm.nih.gov/pmc/articles/PMC7665790/> |  |  |  |
| ***FHS*** | Framingham Heart Study | European Ancestry | Boston University IRB. Boston, MA, USA. | Approved | <https://www.ncbi.nlm.nih.gov/pmc/articles/PMC7665790/> |  |  |  |
| ***GENESTAR 1ea*** | GeneSTAR Study | European Ancestry | Johns Hopkins Medicine IRB. Baltimore, MD, USA. | Approved | <https://www.hopkinsmedicine.org/gim/research/GeneSTAR/index> | <https://www.ncbi.nlm.nih.gov/pmc/articles/PMC7665790/> |  |  |
| ***GENESTAR 1aa*** | GeneSTAR Study | African American | Johns Hopkins Medicine IRB. Baltimore, MD, USA. | Approved | <https://www.hopkinsmedicine.org/gim/research/GeneSTAR/index> | <https://www.ncbi.nlm.nih.gov/pmc/articles/PMC7665790/> |  |  |
| ***GENESTAR 2ea*** | GeneSTAR Study | European Ancestry | Johns Hopkins Medicine IRB. Baltimore, MD, USA. | Approved | <https://www.hopkinsmedicine.org/gim/research/GeneSTAR/index> | <https://www.ncbi.nlm.nih.gov/pmc/articles/PMC7665790/> |  |  |
| ***GENESTAR 2aa*** | GeneSTAR Study | African American | Johns Hopkins Medicine IRB. Baltimore, MD, USA. | Approved | <https://www.hopkinsmedicine.org/gim/research/GeneSTAR/index> | <https://www.ncbi.nlm.nih.gov/pmc/articles/PMC7665790/> |  |  |
| ***GENOA*** | Genetic Epidemiology Network of Arteriopathy study | European Ancestry | Mayo Clinic IRB. Rochester, MN, USA and University of Michigan IRB. Ann Arbor, MI, USA. | Approved | <https://doi.org/10.1001/archinte.164.12.1313> | <https://www.ncbi.nlm.nih.gov/projects/gap/cgi-bin/GetPdf.cgi?id=phd003593.1> |  |  |
| ***GENOA*** | Genetic Epidemiology Network of Arteriopathy study | African American | Mayo Clinic IRB, Rochester, MN, USA, University of Mississippi Medical Center IRB Jackson, MS, USA, and University of Michigan IRB. Ann Arbor, MI, USA. | Approved | <https://doi.org/10.1001/archinte.164.12.1313> | <https://www.ncbi.nlm.nih.gov/projects/gap/cgi-bin/GetPdf.cgi?id=phd003593.1> |  |  |
| ***JHS*** | Jackson Heart Study | African American | Jackson Heart Study Community Ethics Advisory Board. Jackson, MS, USA. | Approved | <https://www.jsums.edu/jsucorc/jhs-community-ethics-advisory-board/> | <https://www.jacksonheartstudy.org/Research> |  |  |
| ***LLS*** | Leiden Longevity Study | European Ancestry | Leiden University Medical Center Ethical Committee. The Netherlands | Approved | <https://doi.org/10.1038/sj.ejhg.5201508> |  |  |  |
| ***MESA*** | Multi-Ethnic Study of Atherosclerosis | European Ancestry | IRBs from Columbia University, New York, Johns Hopkins University, Baltimore, Northwestern University, Chicago, UCLA, Los Angeles, University of Minnesota, Twin Cities, Wake Forest University, Winston Salem in the USA. | Approved | <https://doi.org/10.1093/aje/kwf113> |  |  |  |
| ***MESA*** | Multi-Ethnic Study of Atherosclerosis | African American | IRBs from Columbia University, New York, Johns Hopkins University, Baltimore, Northwestern University, Chicago, UCLA, Los Angeles, University of Minnesota, Twin Cities, Wake Forest University, Winston Salem in the USA. | Approved | <https://doi.org/10.1093/aje/kwf113> |  |  |  |
| ***NELSON*** | Dutch-Belgian Randomized Lung Cancer Screening Trial | European Ancestry | Dutch Minister of Health, and Ethical Committees from University Medical Center Groningen, University Medical Center Utrecht and Kennemer Gasthuis Haarlem in the Netherlands, and University Hospital Gasthuisberg Leuven in Belgium. | Approved | <https://dx.doi.org/10.1102%2F1470-7330.2011.9020> |  |  |  |
| ***RS-I*** | Rotterdam Study I | European Ancestry | Medical Ethics Committee of the Erasmus MC and by the Dutch Ministry of Health, Welfare and Sport. Rotterdam, The Netherlands. | Approved | <https://doi.org/10.1007/s10654-020-00640-5> |  |  |  |
| ***RS-II*** | Rotterdam Study II | European Ancestry | Medical Ethics Committee of the Erasmus MC and by the Dutch Ministry of Health, Welfare and Sport. Rotterdam, The Netherlands. | Approved | <https://doi.org/10.1007/s10654-020-00640-5> |  |  |  |
| ***YFS*** | Young Finns Study | European Ancestry | Ethical Committee of Hospital District of Southwest Finland. Finland. | Approved | <https://doi.org/10.1038/s41598-021-86536-0> |  |  |  |
